# Ethnic inequalities in compulsory psychiatric hospital detentions during UK COVID-19 ‘lockdowns’: A Regression Discontinuity Design in time study

**DOI:** 10.1101/2025.05.02.25326888

**Authors:** R. Hildersley, T.K. Oswald, I Bakolis, L Becares, A. Dregan, J Dyer, M. Hotopf, J. Ocloo, R Stewart, J. Das-Munshi

## Abstract

**Background:** Ethnic inequalities in compulsory psychiatric hospital detentions are well-documented in the UK and internationally. It is unknown how UK COVID-19 ‘lockdown measures’, which led to restrictions in public movement, gatherings, in-person health service delivery, and changes to police powers, further impacted inequalities.

**Aims:** In this study, we assessed whether national ‘lockdown’ measures impacted ethnic inequalities in voluntary and compulsory psychiatric hospital admissions, during the COVID-19 pandemic.

**Methods:** Daily counts of admissions and detentions to psychiatric hospitals were extracted from a large population-level sample of secondary mental health service users in southeast London. Changes during two COVID-19 lockdown periods, over 2020-2021, were compared with pre-pandemic periods (2016-2019) with the use of a regression discontinuity in time design to assess ethnic inequalities in voluntary/ compulsory mental health admissions.

**Results:** Compared to the pre-pandemic reference (2016-2019), after adjusting for seasonal and weekly trends, overall admissions to mental health units dropped during the first COVID-19 lockdown (Incidence Rate Ratio (IRR) 0.87 (95% CI: 0.75-1.00) but with more compulsory detentions (1.25 (1.05-1.54)). This was mostly due to higher compulsory detentions in the Black Caribbean group (1.54 (1.08-2.19)). During the second COVID-19 lockdown, whereas total daily admissions remained similar to the pre-pandemic reference (1.03 (0.92-1.15)), total new daily detentions was elevated (1.28 (1.11-1.49)), specifically in Black Caribbean (1.53 (1.14, 2.06)) and Black African (1.57 (1.06-2.34)) groups.

**Conclusions:** COVID-19 lockdown measures exacerbated pre-existing ethnic inequalities in compulsory psychiatric detention, particularly for those from Black Caribbean and Black African backgrounds. There is a need to address ethnic inequalities in compulsory psychiatric detentions and attend to exacerbations of pre-existing inequalities during health emergencies like the COVID-19 pandemic. This cannot be achieved without addressing systemic racism within criminal justice and healthcare systems and tackling inequalities in wider social and economic determinants of mental health.

**Funding:** Health Foundation, NIHR, UKRI.

## Introduction

The acute phases of the Coronavirus Disease 2019 (COVID-19) pandemic altered most aspects of daily life, impacting the physical and mental health of populations. Attempts to slow the spread of COVID-19 through “lockdown” measures caused disruptions to employment, education, and health service use. Notably, a study of 10 UK mental health service sites reported reduced daily referrals, inpatient admissions, and caseloads during the first UK lockdown period in Spring 2020, compared with a pre-lockdown period, and this observation was also found elsewhere(1, 2) Similarly in the U.S., mental health-related emergency department visits declined from March 2020 relative to a pre-pandemic period, and some evidence suggests there was no significant change in psychiatric admissions.(3, 4)

In the UK, Black and other racially minoritised groups are more likely to come into mental health services through compulsory pathways, frequently involving contact with police and criminal justice,(5, 6) despite no consistent evidence of greater violence or substance misuse.(7) This has also been observed in other countries.(8) During the early stages of the COVID-19 pandemic, police in the UK were given greater powers to enforce pandemic mitigation measures, such as social distancing.(9) It has become evident that there was significant racialised inequality in the way these measures were used.(9)

Further, access to mental health care was not equally distributed during the pandemic, with ethnic inequalities reported in some countries. For example, in the US, compared to White Americans, Black, Hispanic, and Asian Americans were significantly less likely to receive mental healthcare services or telemedicine appointments during the pandemic.(10, 11) Even prior to the pandemic, ethnic inequalities in mental health access and outcomes were known to be a major concern in the UK and internationally.(12)

Although pre-existing ethnic inequalities in compulsory psychiatric hospital detentions are well-documented, a review of the literature (see supplementary material 1) suggests no previous studies have yet investigated how COVID19 lockdowns affected pre-existing ethnic inequalities in mental health-related admissions and detentions. Therefore, using a large population-level sample of people in contact with secondary mental health services in southeast London, the aim of the current study was to investigate (a) how the first and second COVID-19 lockdowns, which occurred in 2020, affected overall mental health hospital admissions (both voluntary and compulsory combined, referred to as “admissions” from this point) and compulsory psychiatric hospital detentions (referred to as “compulsory detentions” from this point), and (b) whether this varied between racialised minority groups, compared to White British mental health service users. The present analysis forms part of a larger mixed methods study which investigated the impact of the COVID19 pandemic on health and service use in racially minoritised people(13)

## Methods

### Participants and setting

Patient-level demographic and hospital admissions data in this study were extracted from mental health medical records from one of Europe’s largest secondary mental healthcare providers, the South London and Maudsley NHS Foundation Trust (SLaM) using its Clinical Record Interactive Search (CRIS) platform.(14) The construction of the dataset is detailed in Figure 1. In the UK, healthcare is free at the point of contact, and, within this context, SLaM is near sole provider of secondary mental health services for 1.3 million residents in a defined geographic area in southeast London. Like other urban areas in the UK, the catchment area for this study is characterised by high ethnicity and socioeconomic diversity (see supplementary materials 1, Table 1 for further details).

**Figure 1:**
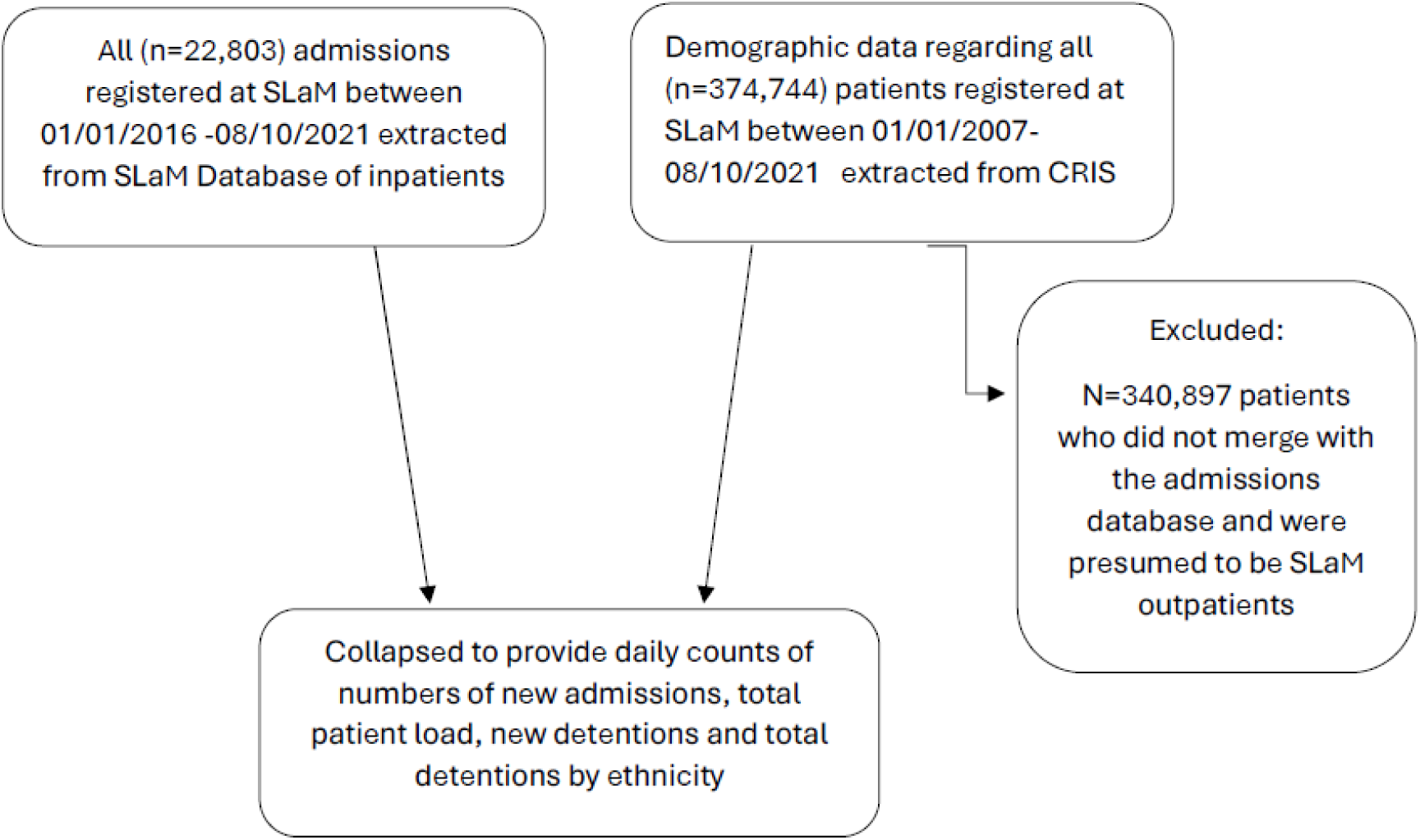
Flow diagram showing construction of dataset for analysis

**Table 1:** Descriptive characteristics (median and interquartile range (IQR)) of the sample, with average number of daily new admissions and detentions by ethnicity.

|  | Full sample |  | 2020 Pre-lockdown -01 Jan 2016 - 22 March 2020 |  | Lockdown 1 - 23 March 2020 - 12 May 2020 |  | Lockdown 1 lift - 13 May 2020 - 1 November 2020 |  | Lockdown 2 - 2 November 2020 - 7 March 2021 |  | Lockdown 2 lift - 8 March 2021 - 8 October 2021 |  |
| --- | --- | --- | --- | --- | --- | --- | --- | --- | --- | --- | --- | --- |
|  | Median | (IQR) | Median | (IQR) | Median | (IQR) | Median | (IQR) | Median | (IQR) | Median | (IQR) |
| Total Daily New Admissions (voluntary admissions + compulsory detentions) |  |  |  |  |  |  |  |  |  |  |  |  |
| Full Sample | 10.00 | (8-13) | 11.00 | (8-13) | 8.00 | (7-10) | 11.00 | (8-13) | 10.00 | (8-12) | 11.00 | (8-14) |
| White British | 3.00 | (2-5) | 3.00 | (2-5) | 2.00 | (1-3.5) | 3.00 | (1-4) | 3.00 | (2-4) | 3.00 | (2-4) |
| White Other | 1.00 | (0-1) | 1.00 | (0-1) | 0.50 | (0-1) | 1.00 | (0-1) | 1.00 | (0-1) | 1.00 | (0-1) |
| Black Caribbean | 2.00 | (1-3) | 2.00 | (1-3) | 2.00 | (1-3) | 2.00 | (1-4) | 2.00 | (1-3) | 2.00 | (1-3) |
| Black African | 1.00 | (0-2) | 1.00 | (0-2) | 1.00 | (0-2) | 1.00 | (0-2) | 1.00 | (1-2) | 1.00 | (0-2) |
| South Asian | 0.00 | (0-0) | 0.00 | (0-0) | 0.00 | (0-0) | 0.00 | (0-0) | 0.00 | (0-0) | 0.00 | (0-1) |
| All other Ethnicities | 1.00 | (0-1) | 1.00 | (0-2) | 0.00 | (0-1) | 1.00 | (0-1) | 1.00 | (0-1) | 1.00 | (0-1) |
| Total Daily New Compulsory Detentions |  |  |  |  |  |  |  |  |  |  |  |  |
| Full Sample | 6.00 | (4-8) | 6.00 | (4-8) | 6.00 | (5-8.5) | 7.00 | (5-9) | 7.00 | (5-9) | 7.00 | (5-9) |
| White British | 1.00 | (0-2) | 1.00 | (0-2) | 1.00 | (0-2) | 1.00 | (.5-3) | 1.00 | (0-2) | 1.00 | (0-2) |
| White Other | 0.00 | (0-1) | 0.00 | (0-1) | 0.00 | (0-1) | 1.00 | (0-1) | 0.00 | (0-1) | 0.00 | (0-1) |
| Black Caribbean | 2.00 | (1-3) | 1.00 | (1-2) | 2.00 | (1-3) | 2.00 | (1-3) | 2.00 | (1-3) | 2.00 | (1-3) |
| Black African | 1.00 | (0-1) | 1.00 | (0-1) | 1.00 | (0-2) | 1.00 | (0-2) | 1.00 | (0-2) | 1.00 | (0-2) |
| South Asian | 0.00 | (0-0) | 0.00 | (0-0) | 0.00 | (0-0) | 0.00 | (0-0) | 0.00 | (0-0) | 0.00 | (0-0) |
| All other Ethnicities | 0.00 | (0-1) | 0.00 | (0-1) | 0.00 | (0-1) | 0.00 | (0-1) | 0.00 | (0-1) | 0.00 | (0-1) |

### Measures

Hospital admissions data was used to provide daily counts of admissions and all classifications of compulsory detentions under the Mental Health Act (1983) (MHA), subdivided by ethnicity. The outcomes of interest were any admission to inpatient psychiatric services over the observation period, either as voluntary patients or as compulsory detentions. The units for analysis used were: (a) the number of *new* admissions/detentions per day (incidence, that is, the number of patients added to the patient load on any given day) and (b) the *total* number of inpatients detained/ all inpatients within in-patient services per day (prevalence, that is, the total patient load compulsorily detained on each given day).

Demographic data, specifically ethnicity, were linked to the full list of admissions to SLaM from 1^st^ January 2016 to 8^th^ October 2021 using pseudonymised unique patient identifiers. Ethnicity was initially mapped on to Census 2011 ethnicity criteria and grouped as: White British, Irish, White Other, Black Caribbean, Black African, Indian, Pakistani, and Bangladeshi, Chinese, Other Ethnicity, Mixed Ethnicity. Due to small sample sizes a decision was taken to aggregate groups, resulting in: White British, White Other (including Irish), Black Caribbean, Black African, South Asian (a combined category including Indian, Pakistani and Bangladeshi groups) and Other Ethnicity (including other mixed ethnicities, Chinese ethnicity and all other Asian ethnicity). Participants of mixed ethnicity were grouped with individuals with their minority ethnic identity, for example, participants who are mixed Black Caribbean and White British were included in the Black Caribbean category.

### Time periods for analysis

England first introduced “lockdown” regulations on 23^rd^ March 2020, which involved closing all “non-essential” businesses, recommending that people work from home if possible, and only allowing people to leave their home for essential purposes (including seeking medical care). These regulations started to be lifted from 13^th^ May 2020, with most national restrictions lifted on 4^th^ July 2020 and replaced with regional regulations. A modified national “lockdown” policy was reintroduced on 5^th^ November 2020 and, while this was replaced by regional regulations on 2^nd^ December 2020, the London regulations remained strict (i.e., limited contact between households, recommendations for people to work from home if possible and closure of “non-essential” businesses). Shortly after this, on 6^th^ January 2021, a third national “lockdown”. was introduced, with a phased exit from lockdown starting on 8^th^ March 2021 and ending on 18^th^ July 2021.(15) These measures were supported by expansions to policing powers, as described above.

For the purpose of this study, we therefore chose to treat the UK lockdown measures as two distinct periods, as the regional restrictions placed on London between the second and third national lockdowns were of the highest level, and similar to those introduced during the national lockdown periods. A detailed timeline of the UK coronavirus mitigation measures can be found here: https://www.instituteforgovernment.org.uk/data-visualisation/timeline-coronavirus-lockdowns. For the purpose of this analysis, the first UK lockdown was defined from 23^rd^ March 2020 to 13^th^ May 2020 and the second from 2^nd^ November 2020 to 8^th^ March 2021. The cut points used for the lockdown periods in the analysis and control periods from 1^st^ January 2016 to 8^th^ October 2017, and 1^st^ January 2018 to 8^th^ October 2019, are illustrated in Figure 2.

**Figure 2:**
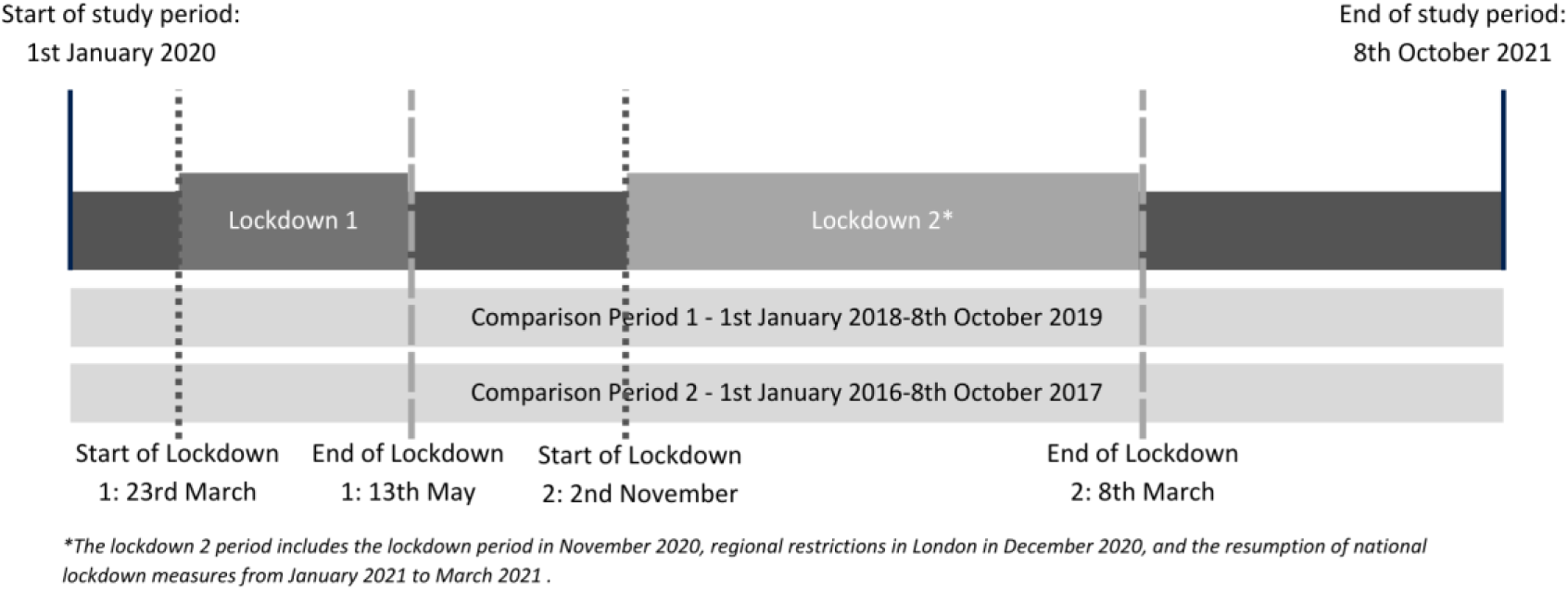
Timelines of the study with cut points and comparison periods. Lockdown periods are sections of time that include regulations relating to the closure of businesses and limited household mixing.

### Statistical methods

First, we calculated the median and interquartile range (IQR) of the number of new and total admissions and detentions for each study period. Next, we used a fuzzy Regression Discontinuity in Time with a Difference in Differences strategy to determine the effect of the four intervention cut points (i.e., the start and lift of the two described “lockdown” periods).(1, 16, 17) We included two parallel pre-pandemic comparison periods in the models (1^st^ January 2016 to 8^th^ October 2017 and 1^st^ January 2018 to 8^th^ October 2019) to account for any changes associated with the intervention time points that were not due to the COVID-19 “lockdown” policies, shown in figure 2. We used Poisson regression models with robust standard errors, to assess the change associated with the cut points, accounting for trends associated with the month and day of the week. Where needed, we ran zero inflated Poisson models to account for the high frequency of zero counts in the models analysing the daily rate of new admissions and compulsory detentions. Diagnostic tests including checking for goodness of fit, autocorrelation, normality of the residuals, and robustness of the linear predictions in the model to ensure assumptions were not breached. First, we assessed the overall change in in the total and newly admitted or detained cases per day associated with the cut points, then we assessed the changes associated with the UK lockdown for the ethnicity subgroups. Further details of the statistical methods, including assumptions, can be found in Supplementary Material 2.

### Post-hoc Analysis

We conducted further post-hoc analyses to assess if observed changes in admissions were due to new admissions of people coming into secondary mental health services during the COVID-19 period for the first time. This post hoc analysis assessed whether potential lower levels of early/ preventative access to community care during the COVID19 pandemic was associated with higher levels of inpatient admissions. To assess this, we re-ran the analyses restricted to people making contact for the first time during the pandemic, i.e., ‘flagged’ as first admissions only, in the clinical data.

### Sensitivity analysis

We ran the following pre-planned sensitivity analyses, as per Bakolis *et al* (2021):(1)

1. To account for anticipatory effects of both lockdown periods potentially impacting people’s behaviours, and UK social distancing measures which were introduced earlier, on 16^th^ March 2020, we re-ran the analysis with the start-of-lockdown dates set to one week before the actual introduction of the policies (i.e., start of lockdown 1 on 16^th^ March 2020 and lockdown 2 on 26^th^ October 2020).
2. To assess the sensitivity of changes associated with the start of the lockdown lifts, we re-ran the analysis with the lockdown ‘lift’ indicators coded as the time periods that had the start of the least restrictive ‘lockdown’ measures (i.e., lift of lockdown 1 set as 4^th^ July 2020 and lockdown 2 as 18^th^ July 2021).
3. To check that the findings were not an artefact of the pre-lockdown time periods, we altered the time window for the lockdown periods to include the period from 30^th^ January to 23^rd^ March 2020 and 28^th^ June 2020 to 2^nd^ November 2020 (i.e., the same number of days as the lockdown period but different specific dates).

### Patient and Public Involvement

Members of our core team had lived experience of the issues under investigation; Dyer has lived experience of caring for siblings with severe mental health issues. The study was part of the COVID-19 Ethnic Inequalities in Mental health and Multimorbidities (COVE-IMM) [project](https://www.kcl.ac.uk/research/covid-19-ethnic-inequalities-in-mental-health-and-multimorbidities-cove-imm-study) which involved an ethnically diverse steering group that included university researchers and people with lived experience of severe mental health problems.(13)

## Results

Daily distributions (medians and IQRs) for the number of new admissions and compulsory detentions for the full sample, grouped by ethnicity, are shown in table 1. The distributions for total number of inpatients and compulsory detentions are displayed in supplementary table 3.1. The median number of admissions per day was 10, with 6 new compulsory detentions per day The median number of inpatients was 697 per day, with 503 compulsorily detained. Overall, total psychiatric admissions and compulsory detentions dropped during the first lockdown period and remained lower than the pre-lockdown period throughout the study period. In contrast, the daily rate of new admissions detained increased over lockdowns compared to the pre-lockdown period and remained higher throughout the study period.

During the first lockdown (23 March 2020 – 12 May 2020), the total number of inpatients admitted, and compulsory detentions fell compared to pre-lockdown trends, illustrated in figure 3a. This trend was consistent for all racially minoritised groups, illustrated in figure 4a. As displayed in Table 2, in adjusted models, the overall number of new admissions reduced slightly (IRR 0.87 (0.75-1.00)) during the first lockdown period and returned to the same rate following the lift of the first lockdown. However, the number of new compulsory detentions rose during the first lockdown (IRR 1.25 (1.02-1.54)). This trend was not noted across all racially minoritised groups, and in fact, was mostly driven by the Black Caribbean group, who experienced a significant rise in daily new detentions compared to the pre-lockdown period (IRR 1.54 (1.08-2.20)) despite not showing a corresponding increase in total new admissions (i.e. comprising both voluntary admissions and compulsorily detentions, IRR 1.17 (0.86-1.58)) Further, there was a consistent decrease in the total number of Black Caribbean patients who were inpatients (IRR 0.81 (0.80-0.28), Supplementary Table 3.2) and total number of Black Caribbean inpatients who were detained (IRR 0.78 (0.77-0.80), suggesting that Black Caribbean patients were being detained more frequently, however were not being kept as inpatients for as long as they had previously.

**Figure 3:**
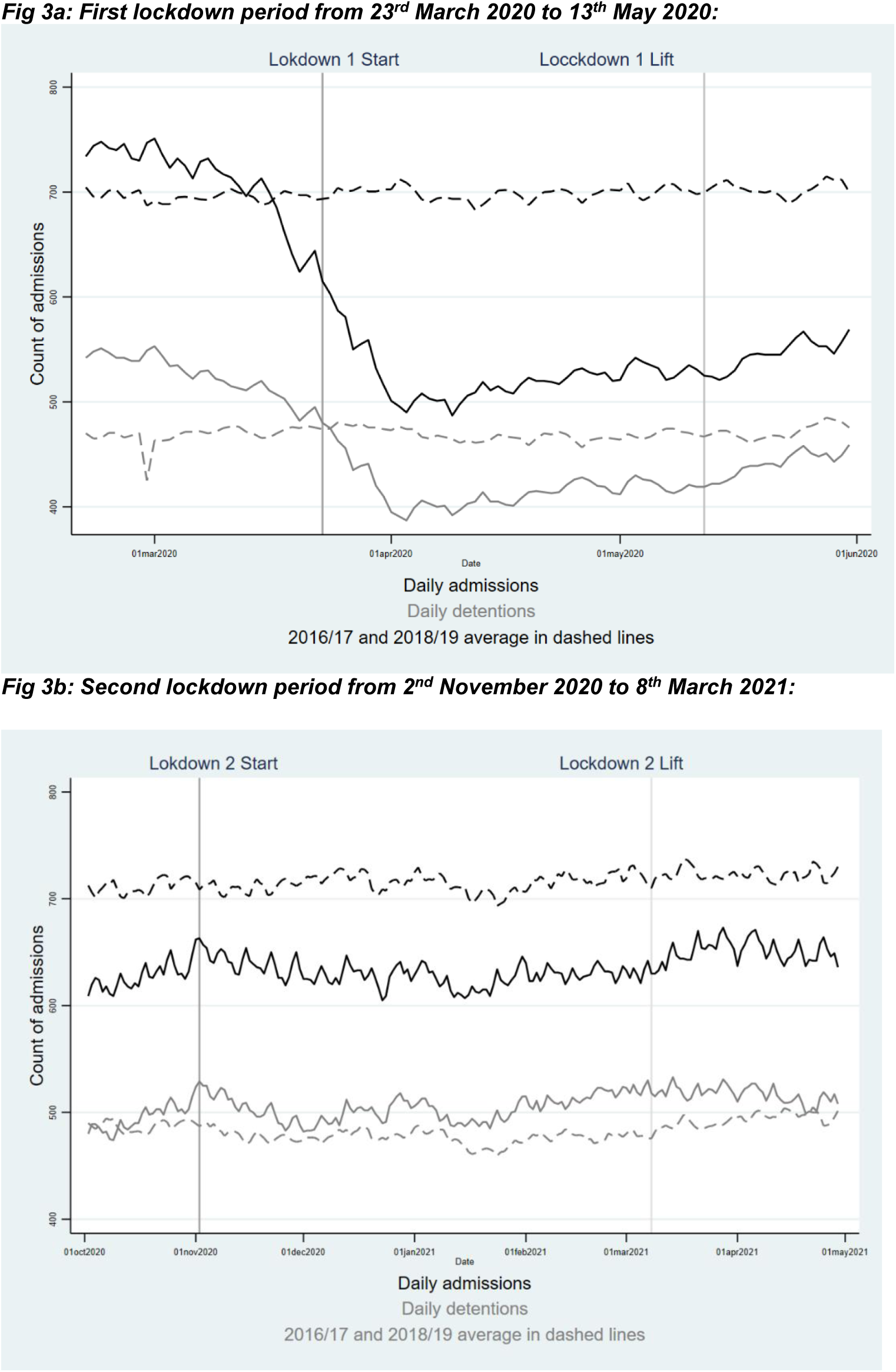
Line graph showing the unadjusted changes in the total number of inpatients and number of inpatients detained within mental health services during (a) the first lockdown period from 23^rd^ March 2020 to 13^th^ May 2020 and (b) the second lockdown period from 2^nd^ November 2020 to 8^th^ March 2021. Dashed lines show the trends during the 2016/2017 and 2018/2019 control periods.

**Figure 4:**
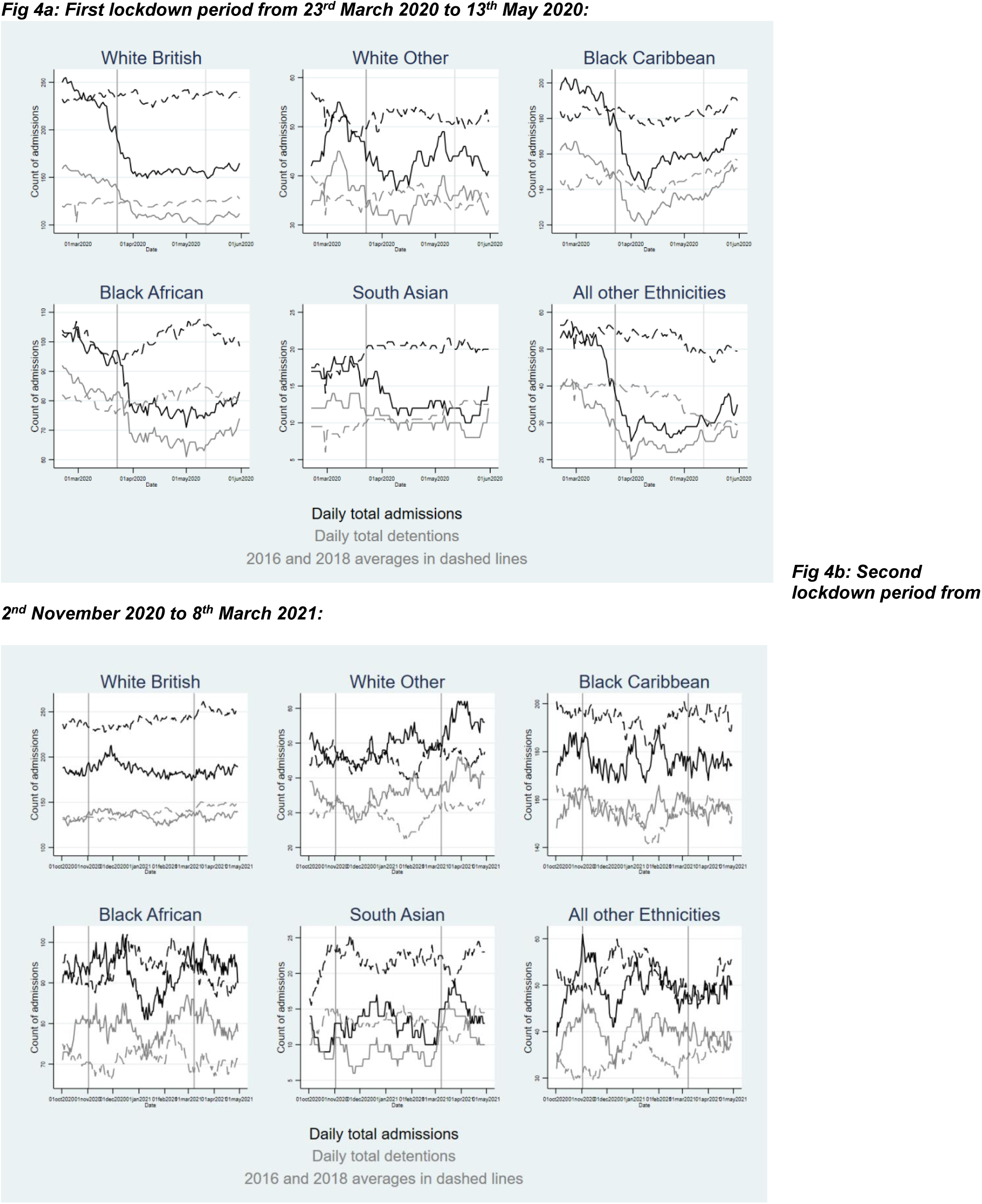
Line graph showing the unadjusted changes in the total number of inpatients and number of inpatients detained, by ethnicity, within SLaM services. Dashed lines show the trends during the 2016/2017 and 2018/2019 control periods.

**Table 2:** Incidence Rate Ratios (IRR) indicating changes in daily admissions (informal admissions + compulsory detentions) and compulsory detentions associated with lockdown compared to the pre-lockdown periods, stratified by ethnicity, adjusted for seasonal and weekly trends. Results obtained using zero inflated Poisson regression models accounting for lower admission rates on different days of the week and robust standard errors.

|  | Lockdown 1 |  |  | Lockdown 1 lift |  |  | Lockdown 2 |  |  | Lockdown 2 lift |  |  |
| --- | --- | --- | --- | --- | --- | --- | --- | --- | --- | --- | --- | --- |
|  | IRR | 95%CI | p-value | IRR | 95%CI | p-value | IRR | 95%CI | p-value | IRR | 95%CI | p-value |
| Daily New Admissions (voluntary admissions + compulsory detentions) |  |  |  |  |  |  |  |  |  |  |  |  |
| Full Sample | 0.87 | (0.75, 1.00) | 0.06 | 1.04 | (0.94, 1.16) | 0.41 | 1.03 | (0.92, 1.15) | 0.57 | 1.14 | (1.03, 1.26) | 0.01 |
| White British | 0.68 | (0.51, 0.89) | 0.01 | 0.97 | (0.79, 1.18) | 0.74 | 0.92 | (0.74, 1.15) | 0.47 | 0.97 | (0.80, 1.18) | 0.75 |
| White Other | 1.08 | (0.62, 1.88) | 0.79 | 1.42 | (0.95, 2.12) | 0.08 | 1.42 | (0.90, 2.23) | 0.13 | 1.67 | (1.11, 2.51) | 0.01 |
| Black Caribbean | 1.17 | (0.86, 1.58) | 0.32 | 1.31 | (1.04, 1.64) | 0.02 | 1.31 | (1.02, 1.67) | 0.03 | 1.26 | (1.01, 1.56) | 0.04 |
| Black African | 0.86 | (0.55, 1.34) | 0.52 | 1.09 | (0.80, 1.49) | 0.56 | 1.28 | (0.92, 1.77) | 0.15 | 1.29 | (0.95, 1.75) | 0.10 |
| South Asian | 1.12 | (0.39, 3.18) | 0.84 | 1.05 | (0.49, 2.25) | 0.89 | 1.15 | (0.53, 2.48) | 0.72 | 1.69 | (0.81, 3.51) | 0.16 |
| All other Ethnicities | 0.57 | (0.34, 0.97) | 0.04 | 0.77 | (0.52, 1.13) | 0.18 | 0.71 | (0.48, 1.05) | 0.09 | 0.87 | (0.60, 1.24) | 0.43 |
| Daily New compulsory detentions |  |  |  |  |  |  |  |  |  |  |  |  |
| Total Daily Admissions Detained | 1.25 | (1.02, 1.54) | 0.03 | 1.16 | (1.01, 1.32) | 0.03 | 1.28 | (1.11, 1.49) | 0.001 | 1.23 | (1.08, 1.40) | 0.001 |
| White British | 0.92 | (0.61, 1.40) | 0.70 | 1.09 | (0.79, 1.49) | 0.61 | 1.01 | (0.72, 1.40) | 0.97 | 0.95 | (0.71, 1.29) | 0.75 |
| White Other | 1.04 | (0.51, 2.13) | 0.90 | 1.30 | (0.75, 2.23) | 0.35 | 1.46 | (0.81, 2.65) | 0.21 | 1.40 | (0.81, 2.41) | 0.23 |
| Black Caribbean | 1.54 | (1.08, 2.20) | 0.02 | 1.34 | (1.03, 1.74) | 0.03 | 1.53 | (1.14, 2.06) | 0.01 | 1.39 | (1.07, 1.79) | 0.01 |
| Black African | 1.14 | (0.69, 1.90) | 0.61 | 1.12 | (0.78, 1.62) | 0.53 | 1.57 | (1.06, 2.34) | 0.03 | 1.35 | (0.94, 1.94) | 0.11 |
| South Asian | 0.50 | (0.12, 2.04) | 0.33 | 0.81 | (0.27, 2.41) | 0.70 | 0.70 | (0.22, 2.19) | 0.54 | 1.22 | (0.41, 3.58) | 0.72 |
| All other Ethnicities | 0.87 | (0.42, 1.80) | 0.71 | 0.90 | (0.53, 1.53) | 0.69 | 0.99 | (0.57, 1.73) | 0.97 | 1.01 | (0.61, 1.68) | 0.97 |

There were fewer changes associated with the second lockdown (2 November 2020 - 8 March 2021), as shown in figure 3b and figure 4b. After adjusting for seasonal and weekly trends, accounting for the control period, there was a slight drop in the total inpatients (IRR 0.87 (0.86-0.27), Supplementary Table 3.2) and compulsory detentions (IRR 0.92 (0.91-0.93), Supplementary Table 3.2) during lockdown 2, and this continued past the lift of the lockdown on 8 March 2021. However, this trend was not universally observed in all racially minoritised groups: the total number of inpatients and compulsory detentions rose for both the White Other and Black African groups in lockdown 2 (supplementary table 3.2). Table 2 shows no overall change in the total number of daily new admissions associated with the second lockdown period, however, there was an increase in the Black Caribbean group (IRR 1.31 (1.02-1.67). Further, there was an increase in the number of new daily compulsory detentions associated with the second lockdown (IRR 1.28 (1.11-1.49)). This appeared to be mainly driven by an increase in new compulsory detentions in the Black Caribbean group (IRR 1.53 (1.14-2.06)) and the Black African group (IRR 1.57 (1.06-2.34)).

In post hoc analyses, we restricted the sample to those who were admitted for the first time to psychiatric units during the pandemic. The restriction was undertaken to assess the possibility that potential changes to community mental health service provision at the time of the pandemic (i.e. reduced access and/ or transition to remote provision over face-to-face contacts), had impacted associations. Despite the sample restriction, which resulted in a smaller overall sample size (initial sample n=15,869 reduced to n=11,033 with restrictions), we found a higher effect size for detentions at all times points in the total sample, but with no changes to estimates for daily new admissions overall. In addition, rates of compulsory admissions were higher in the Black Caribbean group during the second lockdown (IRR 2.07 (95% CI: 1.11, 3.84) in the sample restricted to first contacts vs. IRR: 1.53 (95% CI: 1.14, 2.06) in the unrestricted sample) (supplementary table 5).

In sensitivity analyses we observed evidence of an anticipatory effect in the week leading up to the first UK lockdown, with similar effect sizes to those observed in the main analyses but not as large (see Supplementary Materials 4 Table 4.1). Our findings were robust to further sensitivity analyses which assessed the sensitivity of changes associated with the start of the lockdown lifts (see supplementary table 4.) and sensitivity analyses to check that the findings were not an artefact of pre-lockdown time periods (see supplementary table 4.2).

## Discussion

There have been longstanding concerns that individuals from ethnic minority groups in the UK and across international settings experience inequalities in higher levels of compulsory psychiatric detentions;(8) however, it was unknown how lockdown measures implemented during the COVID-19 pandemic affected this. In line with mental health service trends reported during the early acute stages of the pandemic,(1, 3) we found that admissions to mental health units dropped significantly during lockdown periods for mental health service users within our sample, irrespective of ethnicity. In contrast, there were more detentions across the study sample during lockdown periods, which were driven by higher rates of detentions in Black Caribbean and Black African people relative to pre-lockdown reference periods.

This study is the first to highlight how COVID-19 lockdown measures exacerbated pre-existing ethnic inequalities in compulsory psychiatric detentions. The findings may be attributed to pre-existing social, economic, and health inequalities which were amplified in the pandemic context, alongside concerns that structurally-mediated racism was potentially magnified during the pandemic.(9)

People from some ethnic minority backgrounds in Western countries experience disadvantage relating to social and economic determinants of mental health(18) and policies and laws enacted during the pandemic reinforced and widened these inequalities, having a disproportionate impact on the health and wellbeing of racialised groups.(9, 19) For example, research in the U.S. demonstrated that individuals from ethnic minority groups were more likely to be laid-off or lose employment income during the pandemic,(19, 20) experience housing instability(21) and food insecurity due to financial difficulties.(19, 22) Individuals from ethnic minority groups were also more likely to reside in crowded housing, rely on public transportation, and be working in low-wage essential jobs which lacked paid sick leave, ability to work remotely, and increased regular contact with strangers, directly and indirectly increasing the risk of COVID-19 infection, as well as possible stress and anxiety associated with this.(23) Similar inequalities have been reported in the UK(24, 25). During lockdown periods, restrictions made it difficult for families to cope with COVID-19-related losses, as they could not be involved in the normal processes of death (e.g., saying goodbye or holding a funeral) and access to usual social support systems were reduced.(26, 27)

Outside of the pandemic context, studies from both the UK and US have argued that ethnic minority groups often fear seeking treatment for early symptoms of mental illness due to experiences of racism and discrimination, stigma associated with religious/spiritual practices, mistrust of services, lack of knowledge, fears of confidentiality, and language and cultural barriers.(6, 12, 28–30) This underrepresentation in primary mental health care can lead to more chronic episodes of mental illness, greater functional limitations, and overrepresentation in crisis pathways. During the pandemic, there were challenges in reaching some ethnic minority groups in the UK following closure of community organisations and the transition to remote service delivery for usual check-ups and providing resources.(27) Community service closures during lock-downs may have made people who were already isolated more vulnerable.(27) The provision of remote healthcare would have further disadvantaged those due to the ‘digital divide’ (the gap between those who have access to digital technologies and those who do not).(30). A previous qualitative study of racially minoritised service users from the same catchment of this study, specifically highlighted the adverse impacts of the abrupt withdrawal and/ or rapid transition to telephone and video contacts for the delivery of mental healthcare, hospital appointments, social service and community/ voluntary organisation contacts on the mental health of people living with severe mental health conditions and on their carers (27). Participants in this study reported how the withdrawal of support adversely impacted their health and ability to access care, with resultant detrimental impacts on pathways into care, which were then delayed or disrupted (27). The findings in the study highlighted at least one account of police intervention, which was attributed to disruptions in access to home treatment teams during the pandemic (27). In addition, our post-hoc analyses suggested that when the sample was restricted to first admissions (i.e. excluding those not previously known to mental health services) rates of compulsory detentions were even higher for the total sample. In addition, rates of compulsory detentions were higher for Black Caribbean service users during the second lockdown, in the sample restricted to first contacts. This could imply that changes to UK community health service provision during the pandemic were potentially implicated in higher compulsory admissions as early/ timely access may have been disrupted, however this latter observation will need further exploration.

Some adults from Black ethnic backgrounds in England have described mental health services as disempowering,(31) and detention as a racist and racialised experience, inseparable from a wider context of systemic racism, inequality, and stigma within families and communities.(32) The numbers of people compulsorily detained for treatment in the UK has doubled since 1983 when the most recent legislation governing compulsory admissions to inpatient mental health care was introduced, despite the prevalence of psychosis remaining broadly stable over the same time period.(33) Our observation that there may have been similar, or even elevated rates of compulsory admission in people who had not previously been admitted to mental health services during the pandemic further highlights the potential increase in use of compulsory admissions. In recognition of this increase in detentions, and the persisting ethnic inequalities, there are current moves to reform UK legislation regarding compulsory detention of people suffering from mental health disorders.(34, 35) The review included the development of a Patient and Carers Race Equality Framework that aims to improve mental health services for people who belong to racialised minority groups.(36) While these developments are important in addressing the overrepresentation of ethnic minority individuals in mental health crisis pathways, the present study has shown that we need to ensure that systems are also responsive and effective during times of crisis, in which inequalities across a range of wider determinants of mental health are synergistically exacerbated for individuals from marginalised and ethnic minority groups.(23). In addition, our findings suggest a need to directly tackle systemic racism in criminal justice and healthcare systems. The methods to address this at systems-level may vary across international contexts. An example of an approach includes the rollout across England of the Patient and Carer Race Equality Framework (PCREF), following the 2018 Mental Health Act Review which highlighted high rates of coercive admissions in racialised minority groups (27). PCREF in England aims to embed anti-racism within mental health service provision, through partnership working with racialised communities leading to coproduced services, through interventions to tackle racism and implicit biases in the workforce, and through improving data collection, leading to better monitoring and delivery of equitable mental healthcare for racialised communities. Although this is an important initiative which will potentially improve secondary mental health care in England, previous research has indicated that experiences of racism and racial discrimination are pervasive and persistent across domains, and throughout the life course(37). As such, anti-racism initiatives that target single systems are not sufficient to address structural racism; what is needed are multisector, equity-oriented initiatives and policy reform.(38)

This study’s strength lies in the utilisation of routine data from a large healthcare provider that covers an ethnically diverse population within southeast London. In the UK, most care is provided freely under the National Health Service (NHS), with the mental health Trust in the present study being the sole NHS secondary mental healthcare provider in this area. Therefore, we can be confident that the analyses would have captured most mental health hospital admissions within the catchment, although a few private providers do also operate in the area of the study. Additionally, the study was conducted over a 5-year period spanning a pre-pandemic period which enabled robust comparisons with a prior reference period, and we adjusted for seasonal and weekly trends across models.

Some limitations should be considered. First, the study reports the effect of UK lockdowns on one healthcare provider in one region of the UK. The trends observed in this study may differ from other regions in the UK, however are likely to generalise to other similar urban, and ethnically diverse catchment areas elsewhere. There were also limitations in sample size, particularly for some minority ethnic groups, which may have led to some analyses being underpowered. We had to combine ethnicity groups due to smaller sample sizes, (eg. ‘South Asian’ group included Indian, Pakistani, Bangladeshi people), which would have masked heterogeneity.

Further, due to the study design, we assumed other characteristics, such as area deprivation, remained constant between the different comparison periods and cut points, and we were unable to account for the potential confounding effect of these and other factors within our analysis. Our study only followed the sample during the initial lockdown phases of the pandemic, and so should not be extrapolated out of this context.

In conclusion, there is a need to address ethnic inequalities in compulsory psychiatric detentions and attend to the exacerbation of pre-existing social and economic inequalities during health emergencies like the COVID-19 pandemic, in the UK and internationally. This cannot be achieved without first ensuring primary mental health care is culturally safe and appropriate, reaching ethnic minority groups early. In England, Mental Health Act reform will play a role in potentially preventing these evident ethnic inequalities within inpatient mental health services. Importantly, there is a need to address systemic racism within criminal justice and healthcare systems to reduce the overrepresentation of ethnic minority individuals in crisis pathways.

## Contributorship

JDM, IB, RH conceived and designed the study. JDM led acquisition of the data for analysis. Analysis of the data was led by RH, supported by JDM and IB. All authors-RH, TO, IB, LB, AD, JD, MH, JO, RS, JDM contributed to interpretation of data analyses. Drafting of the work was led by RH and TO, supported by JDM. All authors (RH, TO, IB, LB, AD, JD, MH, JO, RS, JDM) reviewed the work critically for important intellectual content. All authors (RH, TO, IB, LB, AD, JD, MH, JO, RS, JDM) have approved the final version for it to be published. All authors agree to be accountable for the work done.

## Data Availability

The data are not publicly available. Data are owned by a 3rd party SLaM BRC CRIS tool which provides access to anonymised data derived from SLaM electronic medical records. These data can only be accessed by permitted individuals from within a secure firewall (i.e. remote access is not possible and the data cannot be sent elsewhere) in the same manner as the authors, following relevant project approvals.

Analytic code may be available by contacting the lead author (RH) or corresponding author (JDM).

