## Supplementary Materials for "Ethnic inequalities in compulsory psychiatric hospital detentions during UK COVID-19 ‘lockdowns’: A Regression Discontinuity Design in time study"

**Tables:** 5

**Supplementary Material Table 1: Background Information**

Scoping Literature Search

**Evidence before this study**

To assess the evidence for ethnic inequalities in compulsory psychiatric hospital detentions during COVID-19 lockdowns, we systematically searched for primary quantitative studies in Medline (OVID) from 01 January 2020 until 19 September 2024. We used the search term "((‘compulsory detention*’ OR ‘compulsory admission*’ OR ‘involuntary admission*’ OR ‘involuntary treatment’ OR ‘compulsory treatment’ OR ‘psychiatric admission*’ OR ‘psychiatric detention’ OR ‘mental health act’ OR ‘mental health care act’ OR ‘mental health legislation’ OR ‘admission order*’ OR ‘detention order*’) AND (COVID-19  OR coronavirus OR lockdown* OR restriction* OR pandemic) AND (ethnic* OR race OR racial))", with no language restrictions.

Five studies were retrieved and screened. Of these, two studies contained potentially relevant information. One study in Cambridgeshire and Peterborough, UK (population ~860,000), reported that both detained and voluntary inpatient numbers dropped sharply due to the COVID-19 lockdown (from March 2020), and there was no evidence for a change in the use of the Mental Health Act within the mental health service or by police.[1] However, this study did not report findings by patient ethnicity. Another study in the U.S. aimed to characterise and compare inpatient psychiatric admissions in West Texas before (March – July 2019) and during (March – July 2020) the initial months of the COVID-19 pandemic (~800,000 local and surrounding inhabitants combined, with a sizeable refugee population).[2] Voluntary and involuntary psychiatric admissions were combined in analyses and no significant difference in psychiatric admissions was reported between pre- and during-pandemic periods. There was no significant association between race/ ethnicity and the different periods, but there was a significantly higher percentage of patients who identified as Hispanic/Latinx in the during-pandemic period compared to the pre-pandemic period. No studies were identified which specifically investigated ethnic inequalities in compulsory psychiatric hospital detentions during COVID-19 lockdowns.

**Added value of this study**

Our study is the first to investigate ethnic inequalities in compulsory psychiatric hospital detentions during the COVID-19 lockdowns, using longitudinal routine data from a regionally representative healthcare provider that covers a large, ethnically diverse population within southeast London (population ~1.3 million residents). Our findings show that there were more compulsory psychiatric detentions across the study sample during lockdown periods, which were driven by higher rates of

---

[1] Chen S, Jones PB, Underwood BR, Moore A, Bullmore ET, Banerjee S, et al. The early impact of COVID-19 on mental health and community physical health services and their patients’ mortality in Cambridgeshire and Peterborough, UK. Journal of psychiatric research. 2020;131:244-54.

[2] Kim J, Rao N, Collins A, Eboh T, Chugh J, Sheladia S, et al. Retrospective Study of Psychiatric Hospitalizations in a West Texas Mental Health Treatment Facility during the COVID-19 Pandemic. Southern medical journal. 2023;116(2):170

detentions in Black Caribbean and Black African people. This study is the first to highlight how COVID-19 lockdown measures exacerbated pre-existing ethnic inequalities in compulsory psychiatric detentions.

**Implications of all the available evidence**

There have been longstanding concerns that individuals from racialised minority groups in the UK and across international settings experience inequalities in higher levels of compulsory psychiatric detentions. Our findings highlight that these inequalities may be further exacerbated during health emergencies like the COVID-19 pandemic in which laws enacted reinforce and widen social and economic inequalities, having a disproportionate impact on the health and wellbeing of marginalised groups. There is a need to pay close attention to and address the exacerbation of pre-existing inequalities during health emergencies like the COVID-19 pandemic. This cannot be achieved without addressing systemic racism within criminal justice and healthcare systems and tackling inequalities in wider social and economic determinants of mental health.

### Study Catchment Area Demographic details

The study population is drawn from the records of patients in the South London and Maudsley NHS foundation trust, which is near sole provider of secondary mental health care in the London boroughs of Croydon, Lambeth, Lewisham and Southwark, which are four local authorities situated in southeast London, covering a regional catchment area of 1.3 million people.  Supplementary table 1.1 shows a comparison between the differences in social and demographic characteristics of the study catchment area and the rest of England and Wales. The data for this table are drawn from area-level statistics produced by the Office for National Statistics drawn from the 2021 UK Census.

**Supplementary Table 1: Average socioeconomic and sociodemographic composition of local authorities in England and Wales, and in the catchment area for the study**

| | **England and Wales** | | **Study Catchment Area** | |
|---|---|---|---|---|
| | Mean | SD | Mean | SD |
| **% of local authority by ethnicity** | | | | |
| Black African | 2.35% | 2.98 | 12.60% | 2.28 |
| Black Caribbean | 0.89% | 1.34 | 8.70% | 1.99 |
| Mixed | 2.76% | 1.68 | 7.75% | 0.44 |
| White British | 74.59% | 21.82 | 36.92% | 0.96 |
| White Other | 7.41% | 6.01 | 14.67% | 2.76 |
| Asian | 9.38% | 10.69 | 10.93% | 4.51 |
| **Migration** | | | | |
| % of local authority born in the UK | 83.08% | 13.77 | 62.55 | 2.7 |
| **Household deprivation** | | | | |
| % of households in local authority experiencing zero dimension of deprivation | 47.60% | 4.95 | 48.42% | 1.18 |
| % of households in local authority experiencing 1 dimension of deprivation | 33.47% | 1.52 | 32.50% | 0.93 |
| % of households in local authority experiencing 2 dimensions of deprivation | 14.69% | 2.68 | 14.40% | 0.61 |
| % of households in local authority experiencing 3 dimensions of deprivation | 3.99% | 1.33 | 4.25% | 0.51 |
| % of households in local authority experiencing 4 dimensions of deprivation | 0.24% | 0.13 | 0.38% | 0.05 |

**Reference: Office for National Statistics, available at https://www.nomisweb.co.uk/sources/census_2021 (retrieved 31st October 2024)*

**Supplementary Material 2: Regression Discontinuity in Time with a Difference-in-Difference approach**

To model changes in hospitalization associated with the introduction of the COVID-19 lockdown in the UK, we adopted a fuzzy Regression Discontinuity in Time with a Difference in Differences (fRDiT-DiD) approach, as per previous studies using similar datasets.[3] This was appropriate as the intervention (i.e., the introduction of the lockdown) was uniformly introduced for the whole population with clearly defined temporal cut-points. Regression discontinuity designs are also well suited to looking at the immediate periods before and after an intervention. Further, the addition of control time periods from 2016/17 and 2018/19 to include a difference in differences approach to estimation, which effectively allows us to isolate the effect of the intervention within the dataset. Daily counts of admissions were the most appropriate unit of analysis, as person-level analysis introduced the complication of how to deal with multiple admissions during the specified time period or admissions that spanned multiple cut-points. Further, we did not have adequate data or statistical power to calculate the excess or deficit in admissions due to the lockdown policies in this analysis.

First, we visualised the graphical representation of the changes in rates and inpatient load for the whole cohort and the subset of the cohort who were detained by ethnicity on either side of the lockdown periods. We checked distributions of all variables and used correlograms to check for autocorrelation, partial autocorrelation, stationarity and seasonality within the data. We used a Poisson regression model to account for the passage of time modelled within this time series as negative binomial regression models did not fit the data. Where there was a high frequency of zero counts within the data, that is the daily admissions and compulsory detention rates, we used zero inflated Poisson models. We parameterised the model to account for expected lower rates of admissions and compulsory detentions on different days, such as weekends. Further, we used robust standard errors to account for some predicted minor breaches of the modelling assumptions.

We selected the two-year period in 2020-2021 as the intervention period, with the periods from 2016-2017 and 2018-2019 acting as control groups. We created dummy variables to indicate the cut points, in the intervention and control periods. For example, the period from 23rd March – 13th May were coded as 1 in a variable named uk_lockdown (for the first year of the four analysis periods. We fitted an

---

[3] Bakolis, I., Stewart, R., Baldwin, D., Beenstock, J., Bibby, P., Broadbent, M., Cardinal, R., Chen, S., Chinnasamy, K., Cipriani, A., Douglas, S., Horner, P., Jackson, C.A., John, A., Joyce, D.W., Lee, S.C., Lewis, J., McIntosh, A., Nixon, N., Osborn, D., Phiri, P., Rathod, S., Smith, T., Sokal, R., Waller, R., Landau, S., 2021. Changes in daily mental health service use and mortality at the commencement and lifting of COVID-19 'lockdown' policy in 10 UK sites: a regression discontinuity in time design. BMJ Open 11, e049721. https://doi.org/10.1136/bmjopen-2021-049721

Hildersley, R., Easter, A., Bakolis, I., Carson, L., Howard, L.M., 2022. Changes in the identification and management of mental health and domestic abuse among pregnant women during the COVID-19 lockdown: regression discontinuity study. BJPsych open 8, e96. https://doi.org/10.1192/bjo.2022.66

interaction term between the intervention indicator ($X_1$) and control period indicator ($X_2$) to describe the change associated with the intervention while accounting for seasonal changes. The equation below represents the simplified structure of the model. The coefficient $\beta_3$ represents the change associated with the intervention at the different levels, which is the effect that we are most interested in for this study. $X_{3\text{-}6}$ are included in the model to account for seasonal and weekly trends within the time series.

$$Y_t = \beta_0 + \beta_1 X_1 + \beta_2 X_2 + \beta_3 X_1 * X_2 + \beta_4 X_3 + \beta_5 X_4 + \beta_6 X_5 + \beta_7 X_6 + \varepsilon$$

| | | |
|---|---|---|
| $X_1$ | Intervention indicator (Coded as 0 1st Jan Y1-22nd Mar Y1; 1 23rd Mar Y1-13th May Y1; 2 14th May Y1-1st Nov Y1; 3 2nd Nov Y1 – 8th March Y2; 4 9th March Y2 – 8th Oct Y2) | |
| $X_2$ | Control Period (Coded as 0 2016/17; 1 2018/16; 2 (Intervention period) 2020/21) | |
| $X_3$ | Month (1-12) | Included to control for weekly and yearly trends of admission within the data. |
| $X_4$ | Week-day (1-7) | |
| $X_5$ | English Public Holiday (0/1) | |
| $X_6$ | Weekends (0/1) | |
| $\beta_0$ | Constant | |
| ε | Error term | |

We checked for overdispersion, goodness of fit, autocorrelation and normalcy of the residual distributions to ensure that assumptions of the models were met. We chose to not to include a measure of the rate of change between the cut points as the trends were not linear.

**Supplementary Materials 3: Additional Tables**
***Supplementary Table 3.1***

***Descriptive characteristics (median and interquartile range (IQR)) of the sample, with average number of daily new admissions and detentions by ethnicity***

| | Full sample | | 2020 Pre-lockdown -01 Jan 2016 - 22 March 2020 | | Lockdown 1 - 23 March 2020 - 12 May 2020 | | Lockdown 1 lift - 13 May 2020 - 1 November 2020 | | Lockdown 2 - 2 November 2020 - 7 March 2021 | | Lockdown 2 lift - 8 March 2021 - 8 October 2021 | |
|---|---|---|---|---|---|---|---|---|---|---|---|---|
| | Median | (IQR) | Median | (IQR) | Median | (IQR) | Median | (IQR) | Median | (IQR) | Median | (IQR) |
| Total Number of Inpatients (voluntary admissions + compulsory detentions | | | | | | | | | | | | |
| Full Sample | 697.00 | (663-723) | 711.00 | (694-732) | 522.00 | (509.5-532) | 603.00 | (589-616) | 630.00 | (623-637) | 661.00 | (653-669) |
| White British | 235.00 | (197-243) | 239.00 | (233-246) | 156.00 | (154-158.5) | 182.00 | (176.5-188) | 185.00 | (181-192) | 191.00 | (186-197) |
| White Other | 50.00 | (45-56) | 50.00 | (45-54) | 43.00 | (40.5-44) | 50.00 | (47-54) | 48.00 | (45-51) | 59.00 | (56-62) |
| Black Caribbean | 186.00 | (177-196) | 192.00 | (184-199) | 157.50 | (152-159.5) | 170.00 | (164-175) | 175.00 | (172-179) | 178.00 | (174-185) |
| Black African | 94.00 | (89-98) | 95.00 | (90-100) | 77.00 | (75-78.5) | 90.00 | (86-94) | 93.00 | (88-96) | 91.00 | (85-96) |
| South Asian | 18.00 | (15-21) | 20.00 | (17-21) | 12.00 | (12-14) | 13.00 | (12-14) | 13.00 | (12-14) | 15.00 | (14-17) |
| All other Ethnicities | 52.00 | (48-56) | 53.00 | (50-57) | 29.00 | (28-30.5) | 41.50 | (37-44.5) | 50.00 | (48-54) | 50.00 | (46-54) |
| Total Number of Inpatients Detained | | | | | | | | | | | | |
| Full Sample | 503.00 | (460-522) | 502.00 | (454-526) | 414.50 | (405-424.5) | 479.00 | (472-486) | 505.00 | (494-513) | 518.00 | (512-526) |
| White British | 136.00 | (125-148) | 142.00 | (126-151) | 109.00 | (106-112) | 126.00 | (121.5-130) | 136.00 | (132-139) | 133.00 | (128-138) |
| White Other | 36.00 | (32-40) | 34.00 | (31-39) | 34.00 | (32-35.5) | 39.00 | (36-42) | 35.00 | (32-36) | 43.00 | (40-46) |
| Black Caribbean | 155.00 | (145-164) | 156.00 | (144-166) | 134.00 | (127-136) | 148.00 | (145-152) | 156.00 | (153-159) | 158.00 | (155-167) |
| Black African | 76.00 | (71-80) | 76.00 | (70-80) | 67.00 | (65-69) | 75.00 | (72-80) | 80.00 | (77-82) | 78.00 | (75-82) |
| South Asian | 12.00 | (10-13) | 12.00 | (11-14) | 10.00 | (10-11) | 12.00 | (10.5-13) | 9.00 | (8-10) | 12.00 | (11-14) |
| All other Ethnicities | 36.00 | (33-41) | 36.00 | (33-40) | 24.00 | (23-25) | 33.00 | (29-36) | 41.00 | (39-43) | 40.00 | (38-44) |

*Supplementary Table 3.2*

**Incidence Rate Ratios (IRR) with 95% Confidence Intervals (95% CI) indicating changes in the total number of inpatients and inpatients detained, associated with lockdown compared to the pre-lockdown periods, stratified by ethnicity. Results obtained using Poisson regression models with robust standard errors.**

| | **Lockdown 1** | | | **Lockdown 1 lift** | | | **Lockdown 2** | | | **Lockdown 2 lift** | | |
|---|---|---|---|---|---|---|---|---|---|---|---|---|
| | **IRR** | **95%CI** | **p value** | **IRR** | **95%CI** | **p value** | **IRR** | **95%CI** | **p value** | **IRR** | **95%CI** | **p value** |
| **Total inpatients per day (total voluntary admissions + compulsory detentions)** | | | | | | | | | | | | |
| **Full sample** | 0.72 | (0.71, 0.73) | p <0.001 | 0.80 | (0.79, 0.81) | p <0.001 | 0.87 | (0.86, 0.87) | p <0.001 | 0.90 | (0.89, 0.91) | p <0.001 |
| White British | 0.64 | (0.63, 0.65) | p <0.001 | 0.74 | (0.72, 0.75) | p <0.001 | 0.78 | (0.76, 0.79) | p <0.001 | 0.74 | (0.73, 0.75) | p <0.001 |
| White Other | 0.94 | (0.90, 0.97) | 0.001 | 1.00 | (0.97, 1.04) | 0.869 | 1.22 | (1.18, 1.26) | p <0.001 | 1.23 | (1.19, 1.27) | p <0.001 |
| Black Caribbean | 0.81 | (0.80, 0.82) | p <0.001 | 0.83 | (0.82, 0.84) | p <0.001 | 0.88 | (0.87, 0.89) | p <0.001 | 0.92 | (0.91, 0.93) | p <0.001 |
| Black African | 0.78 | (0.76, 0.81) | p <0.001 | 0.91 | (0.90, 0.93) | p <0.001 | 1.04 | (1.03, 1.06) | p <0.001 | 1.10 | (1.08, 1.12) | p <0.001 |
| South Asian | 0.40 | (0.37, 0.43) | p <0.001 | 0.45 | (0.42, 0.48) | p <0.001 | 0.40 | (0.37, 0.43) | p <0.001 | 0.50 | (0.47, 0.53) | p <0.001 |
| All other Ethnicities | 0.58 | (0.56, 0.60) | p <0.001 | 0.73 | (0.71, 0.75) | p <0.001 | 0.82 | (0.80, 0.84) | p <0.001 | 0.83 | (0.81, 0.85) | p <0.001 |
| **Total compulsory detentions per day** | | | | | | | | | | | | |
| **Full sample** | 0.76 | (0.75, 0.77) | p <0.001 | 0.82 | (0.81, 0.82) | p <0.001 | 0.92 | (0.91, 0.93) | p <0.001 | 0.88 | (0.87, 0.89) | p <0.001 |
| White British | 0.68 | (0.66, 0.69) | p <0.001 | 0.69 | (0.68, 0.71) | p <0.001 | 0.74 | (0.73, 0.76) | p <0.001 | 0.63 | (0.62, 0.64) | p <0.001 |
| White Other | 0.94 | (0.90, 0.98) | 0.003 | 0.96 | (0.93, 0.99) | 0.024 | 1.21 | (1.16, 1.26) | p <0.001 | 1.07 | (1.04, 1.11) | p <0.001 |
| Black Caribbean | 0.78 | (0.77, 0.80) | p <0.001 | 0.82 | (0.80, 0.83) | p <0.001 | 0.94 | (0.92, 0.95) | p <0.001 | 0.96 | (0.95, 0.98) | p <0.001 |
| Black African | 0.77 | (0.75, 0.79) | p <0.001 | 0.89 | (0.88, 0.91) | p <0.001 | 1.06 | (1.05, 1.08) | p <0.001 | 1.05 | (1.03, 1.07) | p <0.001 |
| South Asian | 0.45 | (0.42, 0.49) | p <0.001 | 0.42 | (0.40, 0.45) | p <0.001 | 0.27 | (0.26, 0.29) | p <0.001 | 0.35 | (0.33, 0.37) | p <0.001 |
| All other Ethnicities | 0.72 | (0.69, 0.74) | p <0.001 | 0.96 | (0.92, 0.99) | 0.01 | 1.07 | (1.03, 1.10) | p <0.001 | 1.02 | (0.99, 1.05) | 0.176 |

*Reference: pre-lockdown; adjusted for seasonal and weekly trends*

**Supplementary Materials 4: Sensitivity Analysis**
We conducted three sensitivity analyses:

1. To account for anticipatory effects of both lockdown periods, we re-ran the analysis with the start-of-lockdown dates set to one week before the actual introduction of the policies (i.e., start of lockdown 1 on 16th March 2020 and lockdown 2 on 26th October 2020).
2. To assess the sensitivity of changes associated with the start of the lockdown lifts, we re-ran the analysis with the lockdown 'lift' indicators coded as the time periods that had the start of the least restrictive 'lockdown' measures (i.e., lift of lockdown 1 set as 4th July 2020 and lockdown 2 as 18th July 2021)
3. To check that the findings were not an artefact of the pre-lockdown time periods, we altered the time window for the interventions to look at the period from 30th January-23rd March 2020 and 28th June 2020-2nd November 2020 (i.e., the same number of days as the lockdown period.

The results of these analyses are listed in the tables below.

*Supplementary Table 4.1: Sensitivity analyses 1 and 2*

**Incidence Rate Ratios indicating change associated with lockdown, daily data – Lockdown 1 start: 16th March; lockdown 1 lift: 4th July; lockdown 2 start: 26th October; Lockdown 2 lift: 18th July**

| | Lockdown 1 | | | Lockdown 1 lift | | | Lockdown 2 | | | Lockdown 2 lift | | |
|---|---|---|---|---|---|---|---|---|---|---|---|---|
| | **IRR** | **95%CI** | **p value** | **IRR** | **95%CI** | **p value** | **IRR** | **95%CI** | **p value** | **IRR** | **95%CI** | **p value** |
| **Number of new admissions** | | | | | | | | | | | | |
| Total Daily Admissions - White British | 0.77 | (0.61, 0.97) | 0.03 | 0.93 | (0.74, 1.16) | 0.50 | 0.96 | (0.79, 1.17) | 0.70 | 0.86 | (0.68, 1.09) | 0.20 |
| Total Daily Admissions - White Other | 0.98 | (0.62, 1.53) | 0.93 | 1.51 | (0.98, 2.32) | 0.06 | 1.50 | (1.00, 2.24) | 0.05 | 1.17 | (0.70, 1.96) | 0.56 |
| Total Daily Admissions - Black Caribbean | 1.28 | (0.99, 1.65) | 0.06 | 1.41 | (1.10, 1.81) | 0.01 | 1.32 | (1.06, 1.65) | 0.01 | 1.33 | (1.02, 1.75) | 0.04 |
| Total Daily Admissions - Black African | 0.99 | (0.70, 1.41) | 0.96 | 1.15 | (0.82, 1.61) | 0.41 | 1.29 | (0.95, 1.75) | 0.10 | 1.38 | (0.94, 2.03) | 0.10 |
| Total Daily Admissions - South Asian | 1.11 | (0.46, 2.65) | 0.82 | 0.6 | (0.24, 1.45) | 0.26 | 1.15 | (0.54, 2.43) | 0.72 | 1.80 | (0.71, 4.57) | 0.22 |
| Total Daily Admissions - All other Ethnicities | 0.55 | (0.36, 0.86) | 0.01 | 0.74 | (0.48, 1.14) | 0.17 | 0.78 | (0.54, 1.13) | 0.19 | 0.70 | (0.45, 1.06) | 0.09 |
| **Total number of inpatients** | | | | | | | | | | | | |
| Total Number of Inpatients | 0.74 | (0.73, 0.75) | p <0.001 | 0.81 | (0.80, 0.81) | p <0.001 | 0.87 | (0.87, 0.88) | p <0.001 | 0.90 | (0.89, 0.91) | p <0.001 |
| Total Number of Inpatients - White British | 0.67 | (0.66, 0.68) | p <0.001 | 0.73 | (0.72, 0.74) | p <0.001 | 0.73 | (0.72, 0.74) | p <0.001 | 0.76 | (0.75, 0.78) | p <0.001 |
| Total Number of Inpatients - White Other | 0.93 | (0.90, 0.97) | p <0.001 | 1.08 | (1.04, 1.12) | p <0.001 | 1.26 | (1.22, 1.30) | p <0.001 | 1.16 | (1.12, 1.20) | p <0.001 |
| Total Number of Inpatients - Black Caribbean | 0.82 | (0.81, 0.83) | p <0.001 | 0.82 | (0.81, 0.83) | p <0.001 | 0.89 | (0.88, 0.89) | p <0.001 | 0.94 | (0.92, 0.95) | p <0.001 |
| Total Number of Inpatients - Black African | 0.81 | (0.79, 0.83) | p <0.001 | 0.96 | (0.94, 0.97) | p <0.001 | 1.11 | (1.09, 1.12) | p <0.001 | 0.99 | (0.98, 1.01) | 0.38 |
| Total Number of Inpatients - South Asian | 0.40 | (0.37, 0.43) | p <0.001 | 0.45 | (0.42, 0.48) | p <0.001 | 0.40 | (0.37, 0.43) | p <0.001 | 0.56 | (0.52, 0.60) | p <0.001 |
| Total Number of Inpatients - All other Ethnicities | 0.66 | (0.63, 0.68) | p <0.001 | 0.72 | (0.70, 0.74) | p <0.001 | 0.80 | (0.78, 0.82) | p <0.001 | 0.89 | (0.87, 0.91) | p <0.001 |
| **Number of new detentions** | | | | | | | | | | | | |
| Total Daily Admissions Detained | 1.13 | (0.97, 1.32) | 0.12 | 1.12 | (0.97, 1.30) | 0.11 | 1.26 | (1.11, 1.44) | p <0.001 | 1.14 | (0.97, 1.34) | 0.12 |
| Total Daily Admissions Detained - White British | 1.00 | (0.70, 1.43) | 1.00 | 1.03 | (0.73, 1.44) | 0.88 | 1.03 | (0.76, 1.39) | 0.86 | 0.83 | (0.58, 1.20) | 0.33 |
| Total Daily Admissions Detained - White Other | 0.75 | (0.42, 1.35) | 0.34 | 1.42 | (0.77, 2.62) | 0.26 | 1.39 | (0.81, 2.38) | 0.24 | 0.68 | (0.34, 1.37) | 0.28 |
| Total Daily Admissions Detained - Black Caribbean | 1.45 | (1.08, 1.95) | 0.02 | 1.38 | (1.02, 1.86) | 0.03 | 1.50 | (1.15, 1.96) | 0.003 | 1.41 | (1.03, 1.92) | 0.03 |
| Total Daily Admissions Detained - Black African | 1.15 | (0.76, 1.74) | 0.50 | 1.08 | (0.72, 1.60) | 0.72 | 1.45 | (1.01, 2.08) | 0.05 | 1.30 | (0.83, 2.05) | 0.26 |
| Total Daily Admissions Detained - South Asian | 0.51 | (0.15, 1.75) | 0.28 | 0.43 | (0.12, 1.51) | 0.19 | 0.66 | (0.21, 2.09) | 0.48 | 1.02 | (0.26, 3.97) | 0.97 |
| Total Daily Admissions Detained - All other Ethnicities | 0.67 | (0.37, 1.22) | 0.19 | 0.97 | (0.53, 1.76) | 0.91 | 1.02 | (0.61, 1.72) | 0.93 | 0.90 | (0.50, 1.62) | 0.73 |
| **Total of inpatients detained** | | | | | | | | | | | | |
| Total Number of Inpatients Detained | 0.78 | (0.77, 0.79) | p <0.001 | 0.81 | (0.80, 0.81) | p <0.001 | 0.89 | (0.88, 0.90) | p <0.001 | 0.86 | (0.85, 0.87) | p <0.001 |
| Total Number of Inpatients Detained - White British | 0.68 | (0.66, 0.70) | p <0.001 | 0.68 | (0.67, 0.69) | p <0.001 | 0.66 | (0.65, 0.67) | p <0.001 | 0.64 | (0.63, 0.65) | p <0.001 |
| Total Number of Inpatients Detained - White Other | 0.95 | (0.91, 0.99) | 0.01 | 1.00 | (0.97, 1.04) | 0.82 | 1.2 | (1.15, 1.24) | p <0.001 | 0.94 | (0.90, 0.99) | 0.01 |
| Total Number of Inpatients Detained - Black Caribbean | 0.80 | (0.79, 0.81) | p <0.001 | 0.79 | (0.78, 0.80) | p <0.001 | 0.94 | (0.92, 0.95) | p <0.001 | 0.93 | (0.92, 0.95) | p <0.001 |
| Total Number of Inpatients Detained - Black African | 0.80 | (0.78, 0.81) | p <0.001 | 0.91 | (0.90, 0.93) | p <0.001 | 1.08 | (1.06, 1.10) | p <0.001 | 0.95 | (0.93, 0.97) | p <0.001 |
| Total Number of Inpatients Detained - South Asian | 0.40 | (0.37, 0.42) | p <0.001 | 0.46 | (0.43, 0.49) | p <0.001 | 0.29 | (0.28, 0.31) | p <0.001 | 0.39 | (0.36, 0.41) | p <0.001 |
| Total Number of Inpatients Detained - All other Ethnicities | 0.80 | (0.77, 0.83) | p <0.001 | 0.96 | (0.92, 1.00) | 0.04 | 1.01 | (0.98, 1.03) | 0.72 | 1.14 | (1.11, 1.17) | p <0.001 |

*Adjusted for seasonal and weekly trends*

*Supplementary Table 4. 2: Sensitivity analysis 3*

**Incidence Rate Ratios indicating change associated with lockdown, daily data – Lockdown 1: 30th January-23rd March 2020; Lockdown 2: 28th June 2020-2nd November 2020**

| | **Lockdown 1** | | | **Lockdown 1 lift** | | | **Lockdown 2** | |
|---|---|---|---|---|---|---|---|---|
| | **IRR** | **95%CI** | **P-Value** | **IRR** | **95%CI** | **P-Value** | **IRR** | **95%CI** |
| **Number of new admissions** | | | | | | | | |
| Total Daily Admissions - White British | 0.90 | (0.64, 1.25) | 0.51 | 0.72 | (0.53, 0.98) | 0.04 | 0.93 | (0.70, 1.23) |
| Total Daily Admissions - White Other | 1.32 | (0.66, 2.65) | 0.43 | 1.26 | (0.69, 2.29) | 0.45 | 1.91 | (1.07, 3.41) |
| Total Daily Admissions - Black Caribbean | 1.44 | (0.96, 2.16) | 0.08 | 1.49 | (1.01, 2.20) | 0.04 | 1.72 | (1.19, 2.51) |
| Total Daily Admissions - Black African | 0.75 | (0.43, 1.32) | 0.32 | 0.79 | (0.46, 1.35) | 0.38 | 0.94 | (0.56, 1.56) |
| Total Daily Admissions - South Asian | 0.41 | (0.11, 1.57) | 0.20 | 0.95 | (0.27, 3.32) | 0.94 | 0.43 | (0.13, 1.44) |
| Total Daily Admissions - All other Ethnicities | 0.58 | (0.29, 1.15) | 0.12 | 0.42 | (0.22, 0.80) | 0.01 | 0.58 | (0.31, 1.10) |
| **Total number of inpatients** | | | | | | | | |
| Total Number of Inpatients | 0.97 | (0.96, 0.99) | p <0.001 | 0.73 | (0.72, 0.74) | p <0.001 | 0.80 | (0.79, 0.81) |
| Total Number of Inpatients - White British | 0.92 | (0.90, 0.93) | p <0.001 | 0.63 | (0.62, 0.64) | p <0.001 | 0.71 | (0.70, 0.71) |
| Total Number of Inpatients - White Other | 1.08 | (1.04, 1.13) | p <0.001 | 0.95 | (0.92, 0.99) | 0.02 | 1.10 | (1.06, 1.15) |
| Total Number of Inpatients - Black Caribbean | 0.98 | (0.97, 1.00) | 0.02 | 0.82 | (0.81, 0.83) | p <0.001 | 0.82 | (0.81, 0.83) |
| Total Number of Inpatients - Black African | 1.00 | (0.98, 1.02) | 0.84 | 0.79 | (0.77, 0.81) | p <0.001 | 0.96 | (0.94, 0.98) |
| Total Number of Inpatients - South Asian | 0.57 | (0.52, 0.62) | p <0.001 | 0.28 | (0.26, 0.30) | p <0.001 | 0.31 | (0.30, 0.34) |
| Total Number of Inpatients - All other Ethnicities | 0.98 | (0.94, 1.02) | 0.31 | 0.63 | (0.60, 0.66) | p <0.001 | 0.74 | (0.71, 0.77) |
| **Number of new detentions** | | | | | | | | |
| Total Daily Admissions Detained | 0.94 | (0.75, 1.20) | 0.64 | 1.12 | (0.89, 1.41) | 0.35 | 1.14 | (0.92, 1.42) |
| Total Daily Admissions Detained - White British | 0.80 | (0.49, 1.33) | 0.40 | 0.89 | (0.57, 1.40) | 0.62 | 0.93 | (0.61, 1.41) |
| Total Daily Admissions Detained - White Other | 1.88 | (0.70, 5.05) | 0.21 | 1.33 | (0.56, 3.17) | 0.51 | 2.40 | (1.01, 5.70) |
| Total Daily Admissions Detained - Black Caribbean | 1.26 | (0.79, 2.00) | 0.34 | 1.62 | (1.05, 2.50) | 0.03 | 1.61 | (1.05, 2.45) |
| Total Daily Admissions Detained - Black African | 0.73 | (0.38, 1.40) | 0.34 | 0.93 | (0.51, 1.70) | 0.82 | 0.90 | (0.50, 1.60) |
| Total Daily Admissions Detained - South Asian | 0.29 | (0.02, 3.50) | 0.33 | 0.34 | (0.03, 3.59) | 0.37 | 0.25 | (0.02, 2.55) |
| Total Daily Admissions Detained - All other Ethnicities | 0.60 | (0.23, 1.51) | 0.28 | 0.49 | (0.21, 1.11) | 0.09 | 0.80 | (0.35, 1.81) |
| **Total of inpatients detained** | | | | | | | | |
| Total Number of Inpatients Detained | 0.95 | (0.94, 0.96) | p <0.001 | 0.76 | (0.75, 0.77) | p <0.001 | 0.79 | (0.78, 0.80) |
| Total Number of Inpatients Detained - White British | 0.91 | (0.89, 0.93) | p <0.001 | 0.65 | (0.63, 0.66) | p <0.001 | 0.65 | (0.64, 0.66) |
| Total Number of Inpatients Detained - White Other | 1.08 | (1.03, 1.13) | 0.001 | 0.97 | (0.93, 1.01) | 0.13 | 1.03 | (0.99, 1.07) |
| Total Number of Inpatients Detained - Black Caribbean | 0.93 | (0.91, 0.95) | p <0.001 | 0.77 | (0.76, 0.78) | p <0.001 | 0.77 | (0.76, 0.78) |
| Total Number of Inpatients Detained - Black African | 0.97 | (0.94, 0.99) | 0.01 | 0.77 | (0.75, 0.79) | p <0.001 | 0.91 | (0.89, 0.93) |
| Total Number of Inpatients Detained - South Asian | 0.79 | (0.74, 0.84) | p <0.001 | 0.33 | (0.31, 0.35) | p <0.001 | 0.39 | (0.37, 0.41) |
| Total Number of Inpatients Detained - All other Ethnicities | 0.98 | (0.95, 1.02) | 0.33 | 0.78 | (0.75, 0.80) | p <0.001 | 0.98 | (0.94, 1.01) |

*Adjusted for seasonal and weekly trends*

**Supplementary Materials Table 5: Post-hoc Analyses**

Analyses restricted to new admissions during the COVID-19 pandemic, zero inflated Poisson regression

| | Lockdown 1 | | | Lockdown 1 lift | | | Lockdown 2 | | | Lockdown 2 lift | | |
|---|---|---|---|---|---|---|---|---|---|---|---|---|
| | IRR | 95%CI | p-Value | IRR | 95%CI | p-value | IRR | 95%CI | P-Value | IRR | 95%CI | P-Value |
| Daily New Admissions (voluntary admissions + compulsory detentions) | | | | | | | | | | | | |
| Full Sample | 0.90 | (0.71, 1.15) | 0.41 | 1.04 | (0.88, 1.25) | 0.63 | 1.05 | (0.87, 1.27) | 0.62 | 1.12 | (0.95, 1.33) | 0.18 |
| White British | 0.73 | (0.48, 1.09) | 0.13 | 0.92 | (0.67, 1.25) | 0.59 | 1.00 | (0.73, 1.37) | 0.99 | 0.96 | (0.71, 1.30) | 0.78 |
| White Other | 1.69 | (0.76, 3.73) | 0.20 | 1.25 | (0.69, 2.25) | 0.46 | 1.55 | (0.81, 2.98) | 0.18 | 1.47 | (0.82, 2.63) | 0.20 |
| Black Caribbean | 1.07 | (0.58, 1.99) | 0.82 | 1.23 | (0.78, 1.94) | 0.38 | 1.44 | (0.88, 2.34) | 0.15 | 1.17 | (0.75, 1.83) | 0.49 |
| Black African | 0.87 | (0.42, 1.82) | 0.71 | 1.37 | (0.75, 2.51) | 0.30 | 1.20 | (0.65, 2.22) | 0.56 | 1.01 | (0.56, 1.82) | 0.98 |
| South Asian | 0.81 | (0.15, 4.19) | 0.80 | 0.8 | (0.27, 2.32) | 0.68 | 0.98 | (0.32, 3.04) | 0.97 | 1.04 | (0.36, 3.06) | 0.94 |
| All other Ethnicities | 0.95 | (0.42, 2.14) | 0.91 | 0.96 | (0.55, 1.68) | 0.89 | 0.94 | (0.53, 1.67) | 0.83 | 1.31 | (0.76, 2.24) | 0.33 |
| Daily New compulsory detentions | | | | | | | | | | | | |
| Total Daily Admissions Detained | 1.46 | (1.03, 2.07) | 0.04 | 1.24 | (0.97, 1.59) | 0.09 | 1.45 | (1.10, 1.89) | 0.01 | 1.30 | (1.02, 1.65) | 0.03 |
| White British | 1.35 | (0.72, 2.51) | 0.35 | 1.21 | (0.74, 1.98) | 0.44 | 1.15 | (0.68, 1.96) | 0.60 | 1.12 | (0.69, 1.81) | 0.65 |
| White Other | 2.02 | (0.68, 5.99) | 0.21 | 1.19 | (0.52, 2.71) | 0.68 | 1.62 | (0.66, 4.01) | 0.29 | 1.43 | (0.63, 3.24) | 0.40 |
| Black Caribbean | 1.25 | (0.56, 2.79) | 0.59 | 1.12 | (0.63, 1.97) | 0.70 | 2.07 | (1.11, 3.84) | 0.02 | 1.63 | (0.93, 2.84) | 0.09 |
| Black African | 1.00 | (0.42, 2.38) | 1.00 | 1.43 | (0.67, 3.04) | 0.36 | 1.50 | (0.70, 3.21) | 0.29 | 1.00 | (0.47, 2.11) | 1.00 |
| South Asian | 0.16 | (0.01, 1.84) | 0.14 | 0.69 | (0.17, 2.78) | 0.60 | 0.72 | (0.15, 3.39) | 0.68 | 0.70 | (0.17, 2.97) | 0.63 |
| All other Ethnicities | 1.64 | (0.51, 5.27) | 0.40 | 1.46 | (0.60, 3.54) | 0.40 | 1.57 | (0.63, 3.87) | 0.33 | 1.82 | (0.77, 4.27) | 0.17 |
